# Novel and Known Genetic Players in Hypertension: From Gene Expression to Striking Insights

**DOI:** 10.1101/2025.07.01.25330624

**Authors:** Aiman Farzeen, Riccardo Berutti, Martin Pavlov, Sapna Sharma, Markéta Fuchs, Nadine Lindemann, Birgit Linkohr, eQTLGen Consortium, Moritz von Scheidt, Heribert Schunkert, Annette Peters, Harald Grallert, Melanie Waldenberger, Juliane Winkelmann, Lars Mägdefessel, Alexander Teumer, Holger Prokisch, Christian Gieger

## Abstract

Cardiovascular diseases (CVDs) are the most common non-communicable diseases, responsible for 17.9 million deaths each year. Hypertension is a major risk factor for CVDs. Gene expression data obtained from RNA sequencing can help identify novel genes associated with hypertension and provide deeper insights into the functional roles of the ∼2,000 known hypertension GWAS loci. In this study, we utilized RNA-seq data from 1,796 individuals from the KORA FF4 cohort. Differential gene expression analyses were done for hypertension and its phenotypes, i.e., systolic blood pressure, diastolic blood pressure and pulse pressure, followed by gene set enrichment analyses. The significant genes were then further tested in a sensitivity analysis using participants without any reported cardiometabolic disorders or medication use. To investigate the connection between genetic variants, environment and gene expression, we integrated RNA-seq data, genotyping array data and methylation data to perform eQTL and CpG methylation-gene expression analyses. Mendelian randomization analyses were performed to answer the question of causality between the gene expression of these genes and blood pressure phenotypes. Our findings highlight novel and known genetic contributors to hypertension, establish potential downstream consequences of hypertension-related genetic variants and CpG sites, and reveal that blood pressure is mostly a cause for change in gene expression. We propose 7 novel genes that can be further targeted as candidate genes for functional follow-ups to get insight into potential new pathways for hypertension, i.e.*, MIR23AHG* (also known as *LOC284454*)*, PGPEP1, RNASEK-C17orf49, PDCD4-AS1, SNRPA1, AGO4* and *CCT6P1.* In light of Mendelian randomization analyses and what genes are druggable in our analyses, we recommend *LMNA,* already known to be involved in cardiovascular diseases, as a drug target for managing the adverse effects of hypertension. Our results provide a foundation for future functional studies and drug development efforts.

## Introduction

Blood pressure is the force exerted by circulating blood against the walls of the body’s arteries. It is recorded as two measurements: systolic blood pressure (SBP), which indicates the pressure in the arteries when the heart contracts, and diastolic blood pressure (DBP), which represents the pressure when the heart rests between beats. Another commonly studied phenotype in blood pressure research is pulse pressure (PP), defined as the difference between SBP and DBP and serves as a measure of arterial stiffness. Hypertension (HTN) is defined as elevated SBP ≥ 140 mmHg and/or elevated DBP ≥ 90 mmHg, or controlled blood pressure with antihypertensive medication^1^. Hypertension is the predominant risk factor for cardiovascular diseases (CVDs), and presents a huge burden on the healthcare system around the globe as it affects over 30% of the adult population worldwide^2,3^. A study examining behavioral, environmental, occupational, and metabolic risk factors identified high SBP as the leading contributor of early death globally, and results in approximately 10.8 million preventable deaths each year^4^. Despite advances in our understanding of hypertension, the global incidence of hypertension has increased over the past four decades, warranting further research ^2^.

Hypertension is a multifactorial disorder influenced by genetics, environment and lifestyle. Environmental risk factors for hypertension include stress, high salt intake, lack of physical activity, and obesity^5^. Genetically, hypertension is associated with numerous common genetic variants as well as rare single-gene mutations that affect proteins involved in sodium and water reabsorption^5^. Genetic factors are estimated to account for 30–60% of an individual’s risk of developing hypertension^6^. Evidence for a genetic contribution to hypertension is strengthened by findings such as individuals with a parental or grandparental history of hypertension having a 2.1-fold and 1.33-fold increased risk, respectively, even in the absence of multiple environmental risk factors^7,8^. Additionally, studies have shown that blood pressure is more strongly correlated in monozygotic twins compared to dizygotic twins^9^, further underscoring the heritable nature of the condition. In line with this, genome-wide association studies (GWAS) have identified over 2,000 loci associated with hypertension^10^. However, interpreting non-coding variants and linking them to causal genes and biological functions remains a significant challenge^11–13^. This is where RNA sequencing becomes valuable, as it provides functional insights by quantifying gene expression levels.

In this study, we conducted differential expression analysis by generating and analysing whole-blood RNA sequencing (RNA-seq) data from 1,796 individuals from the KORA FF4 cohort^14^. KORA is a population-based, cross-sectional survey conducted in Augsburg, Southern Germany, in 2013/2014. The FF4 cohort is the second follow-up to the KORA S4 health survey. We used RNA sequencing because it offers higher sensitivity and with that the opportunity to detect new differentially expressed genes (DEGs) compared to array technologies^15–18^. In the literature, more than 30 genes have been reported to be differentially expressed in blood for hypertension. Commonly reported genes for hypertension are *CRIP1, MYADM, TIPARP, TSC22D3, CEBPA, F12, LMNA, TPPP3, FOS, PP1R15A, TAGAP, S100A10, and FGBP2*^19–22^. We performed differential gene expression analyses followed by gene set enrichment analyses. Additionally, we conducted eQTL and CpG methylation-gene expression analyses on the DEGs. This was done to possibly account for the expression of DEGs, taking the genetic and environmental influences into consideration, and to compare which genes experience changes in gene expression from known single nucleotide polymorphisms (SNPs) and 5’—C— phosphate—G—3’ (CpG) sites for hypertension. Lastly, by integrating summary statistics from eQTLGen phase 2, where a genome-wide eQTL analysis was done across 52 cohorts encompassing 43,301 individuals, we conducted Mendelian randomization (MR) analysis to assess the global causality pattern, i.e., whether BP is affected by blood gene expression changes or vice versa. Results from the analyses above shed light on underlying biology behind hypertension and offer new avenues for further functional studies, preventative and therapeutic research.

## Results

### First Phase - Gene Discovery Differential Expression Analysis

Within our analysis, we analysed RNA-seq data from 1,796 individuals and tested 10,498 genes. A detailed overview of the overall characteristics and characteristics stratified by categorical hypertension can be seen in Table 1. We found 5, 56, 40 and 29 differentially expressed genes (p<5×10^−6^) for HTN, SBP, DBP and PP, respectively (see supplementary Table 2). Only 2 genes were common across all four phenotypes analysed, i.e., *MYADM* and *TSPAN2;* these are known to be linked to hypertension in the literature. 71 genes are common in at least two hypertension phenotypes (see Figure 1). We also performed differential expression analysis for women (n = 930) and men (n = 866) separately (see supplementary Table 3). Most of the genes reported in the sex-specific analyses were also reported in the analysis utilizing all individuals, with the exception of some unique ones that are coloured in Figure 1. The differential expression analyses yielded 91 unique genes that we will refer to as the 91 BP-DEGs in the subsequent paragraphs.

**Figure 1:**
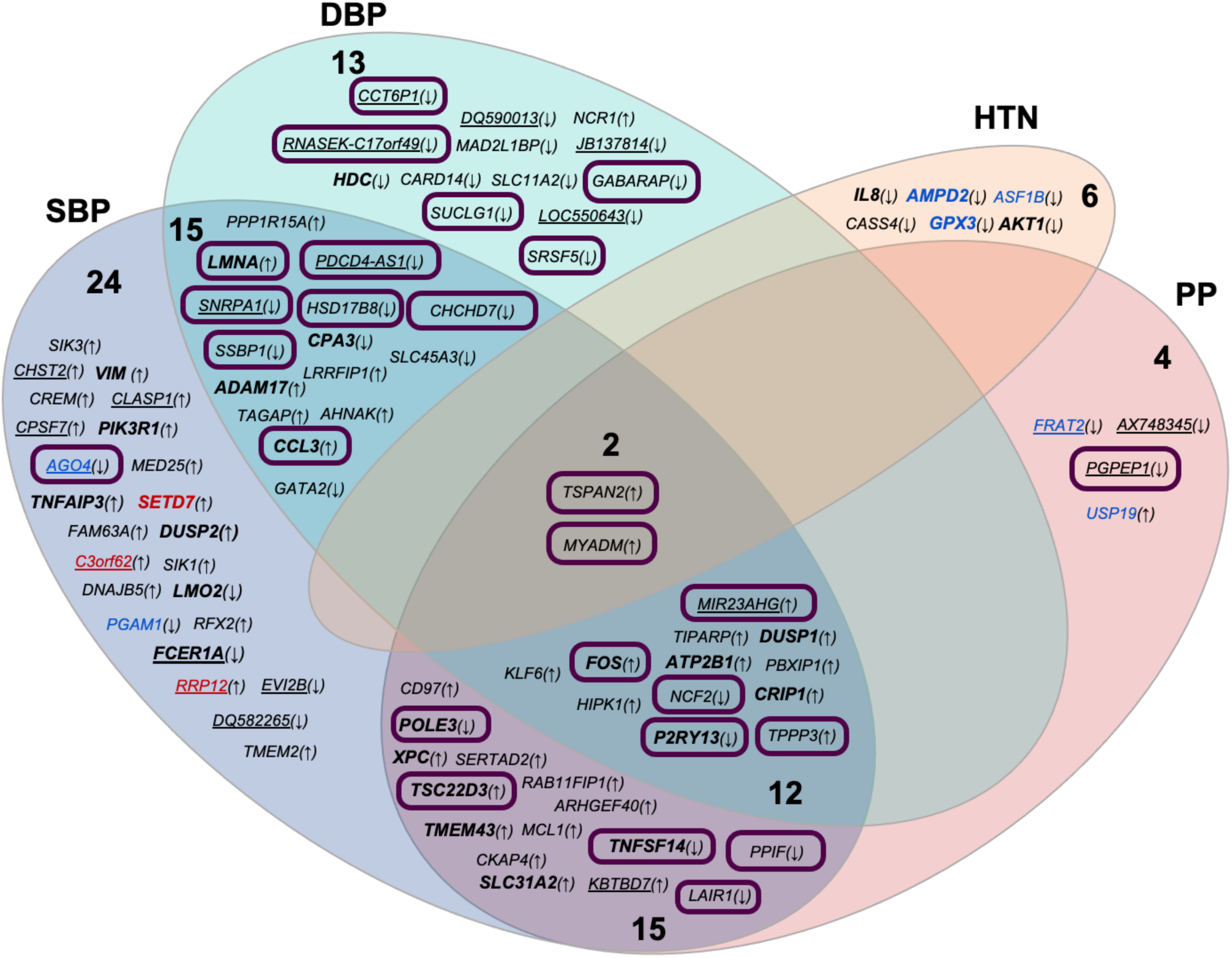
Venn diagram summarizing results from phase 1. Arrows indicate whether a gene is upregulated or downregulated. Genes colored red and blue are specific to women and men, respectively. Enclosed genes are also significant in the sensitivity analysis. Genes in bold are part of the druggable genome. Underlined genes are genes we found to be novel.

**Table 1:** Characteristics of 1,796 study participants stratified by categorical hypertension.

| Summary of continuous variables for the 1,796 individuals |  |  |  |  |  |  |
| --- | --- | --- | --- | --- | --- | --- |
|  | Overall<br>n = 1796 |  | Hypertension = Yes<br>n = 685 |  | Hypertension = No<br>n = 1111 |  |
| Variables | mean | SD | mean | SD | mean | SD |
| Age at checkup (yrs) | 60.0 | 12.3 | 66.7 | 10.6 | 55.9 | 11.5 |
| BMI (kg/m2) | 27.7 | 4.9 | 29.6 | 5.3 | 26.5 | 4.3 |
| Systolic blood pressure (mmHg) | 118.7 | 17.6 | 127.8 | 20.4 | 113.1 | 12.8 |
| Diastolic blood pressure(mmHg) | 72.9 | 9.8 | 75.1 | 12.1 | 71.5 | 7.7 |

| Summary of categorical variables for the 1,796 individuals |  |  |  |  |  |  |  |
| --- | --- | --- | --- | --- | --- | --- | --- |
|  |  | Overall |  | Hypertension = Yes |  | Hypertension = No |  |
| Variable | Level | n | % | n | % | n | % |
| Sex | Male | 866 | 48.2 | 370 | 54 | 496 | 44.6 |
|  | Female | 930 | 51.8 | 315 | 46 | 615 | 55.4 |
| Categorical BMI | Underweight | 10 | 0.6 | 2 | 0.3 | 8 | 0.7 |
|  | Normal weight | 543 | 30.2 | 117 | 17.1 | 426 | 38.3 |
|  | Overweight | 757 | 42.1 | 288 | 42 | 469 | 42.2 |
|  | Obesity Class 1 | 345 | 19.2 | 186 | 27.2 | 159 | 14.3 |
|  | Obesity Class 2 | 96 | 5.3 | 59 | 8.6 | 37 | 3.3 |
|  | Obesity Class 3 | 45 | 2.5 | 33 | 4.8 | 12 | 1.1 |
| Self-reported Angina Pectoris | Yes | 100 | 5.6 | 53 | 7.8 | 47 | 4.2 |
|  | No | 1692 | 94.4 | 630 | 92.2 | 1062 | 95.8 |
| Self-reported Myocardial Infarction | Yes | 63 | 3.5 | 50 | 7.3 | 13 | 1.2 |
|  | No | 1729 | 96.5 | 634 | 92.7 | 1095 | 98.8 |
| Self-reported Stroke | Yes | 51 | 2.8 | 38 | 5.6 | 13 | 1.2 |
|  | No | 1743 | 97.2 | 645 | 94.4 | 1098 | 98.8 |
| Self-reported Cancer | Yes | 192 | 10.7 | 94 | 13.7 | 98 | 8.8 |
|  | No | 1604 | 89.3 | 591 | 86.3 | 1013 | 91.2 |
| Diabetes | No | 1187 | 66.1 | 317 | 46.3 | 870 | 78.3 |
|  | Yes | 609 | 33.9 | 368 | 53.7 | 241 | 21.7 |
| Anti-hypertension Medication | Yes | 622 | 34.6 | 580 | 84.7 | 42 | 3.8 |
|  | No | 1174 | 65.4 | 105 | 15.3 | 1069 | 96.2 |

### Sensitivity Analysis

Given that the study population has a mean age of 60, participants are likely to exhibit age-related ailments—such as cancer or cardiovascular diseases—and may also be on medications to manage these conditions. Hence, we aimed to identify which genes remained significant even after excluding participants who reported having cancer or any cardiometabolic conditions—such as angina pectoris, myocardial infarction, stroke, or diabetes—or were taking medications including lipid-lowering agents, anti-arrhythmics, heart glycosides, antihypertensives, anticoagulants, antithrombotics, oral antidiabetic drugs, or insulin. This was done to have more confidence that the differentially expressed genes were specific for hypertension and not for other cardiometabolic phenotypes or medication. We could include 758 participants in the sensitivity analysis (with HTN n = 51, without HTN = 707). To compensate for the reduced power in the sensitivity analysis and multiple testing, we limited the analyses to the 91 BP-DEGs identified in our differential expression analyses. We found that 19, 17 and 10 genes were significant for SBP, DBP and PP, respectively (see supplementary Table 4). The significant genes from the sensitivity analyses are enclosed in Figure 1.

### Novel and Known Genes: Assessing Druggability and Tissue-Specific Expression

Literature search revealed that 21 genes have not been reported for hypertension before, others are reported for hypertension, other hypertensive conditions such as pulmonary arterial hypertension (PAH) and preeclampsia, or cardiovascular diseases (results summarized in Table 2). Further information on what precisely the genes are known for, along with the references, could be found in the supplementary file.

**Table 2:** Summary of literature search for the 91 BP-DEGs. Arrows indicate whether a gene is upregulated or downregulated in our differential expression analyses. Genes in red are inhibited by drugs in DGIdb, blue genes are both inhibited and activated, and yellow genes are drug targets with no interaction details in DBIdb.

| Comparison of results against published literature | 91 BP- DEGs | Examples |
| --- | --- | --- |
| Reported for hypertension | 30 | *MYADM*(↑), *CD97*(↑), *HIPK1*(↑), *TPPP3*(↑), *ATP2B1*(↑), *TAGAP*(↑), *MCL1*(↑), *TSPAN2*(↑), *LMNA*(↑), *CRIP1*(↑), *KLF6*(↑), *RFX2*(↑), *VIM*(↑), *PPP1R15A*(↑), *AHNAK*(↑), *DUSP1*(↑), *TIPARP*(↑), *SLC31A2*(↑), *DNAJB5*(↑), *TMEM43*(↑), *RAB11FIP1*(↑), *ARHGEF40*(↑), *TSC22D3*(↑), *FAM63A*(↑), *LMO2*(↓), *HSD17B8*(↓), *SLC45A3*(↓), *IL8*(↓), *TNFSF14*(↓), *FOS*(↑) |
| Reported for hypertension in mouse models | 10 | *AKT1*(↓), *DUSP2*(↑), *TNFAIP3*(↑), *PPIF*(↓), *GABARAP*(↓), *NCR1*(↑), *SUCLG1*(↓), *SRSF5*(↓), *SIK1*(↑), *SIK3*(↑) |
| Reported for preeclampsia | 2 | *LAIR1*(↓), *XPC*(↑) |
| Reported for pulmonary arterial hypertension | 2 | *MAD2L1BP*(↓), *GATA2*(↓) |
| Reported for cardiovascular diseases | 16 | *TMEM2*(↑), *CREM*(↑), *P2RY13*(↓), *USP19*(↑), *ASF1B*(↓), *GPX3*(↓), *LRRFIP1*(↑), *PBXIP1*(↑), *SETD7*(↑), *CPA3*(↓), *HDC*(↓), *MED25*(↑), *POLE3*(↓), *CKAP4*(↑), *AMPD2*(↓), *SSBP1*(↓) |
| Protein of the genes reported for hypertension | 4 | *PGAM1*(↓), *ADAM17*(↑), *CASS4*(↓), *CCL3*(↑) |
| SNPs in the genes reported for hypertension | 6 | *CARD14*(↓), *SLC11A2*(↓), *SERTAD2*(↑), *PIK3R1*(↑), *CHCHD7*(↓), *NCF2*(↓) |
| Novel genes for hypertension | 21 | *CLASP1*(↑), *JB137814*(↓), *SNRPA1*(↓), *MIR23AHG*(↑), *AGO4*(↓), *PGPEP1*(↓), *FRAT2*(↓), *C3orf62*(↑), *RRP12*(↑), *PDCD4-AS1*(↓), *RNASEK-C17orf49*(↓), *LOC550643*(↓), *DQ590013*(↓), *CCT6P1*(↓), *KBTBD7*(↑), *DQ582265*(↓), *CPSF7*(↑), *FCER1A*(↓), *EVI2B*(↓), *CHST2*(↑), *AX748345*(↓) |

27 out of the 91 genes are reported as drug targets in the Drug-Gene Interaction Database (DGIdb)^23^, most are reported to be inhibited by drugs in DGIdb, only *ATP2B1* and *AMPD2* display both inhibition and activation interaction with the drugs. 9 genes are drug targets, but their interaction type is classified as unknown in the DGIdb. Among the novel genes, only *FCER1A* was druggable. Additionally, we report the tissues in which these genes show the highest expression, using the Human Protein Atlas^24^ and GTEX^25^ (see Figure 2). Several genes that we detected in whole blood showed their highest expression in bone marrow, lymphoid tissue and brain, exhibiting involvement of multiple organs in hypertension

**Figure 2:**
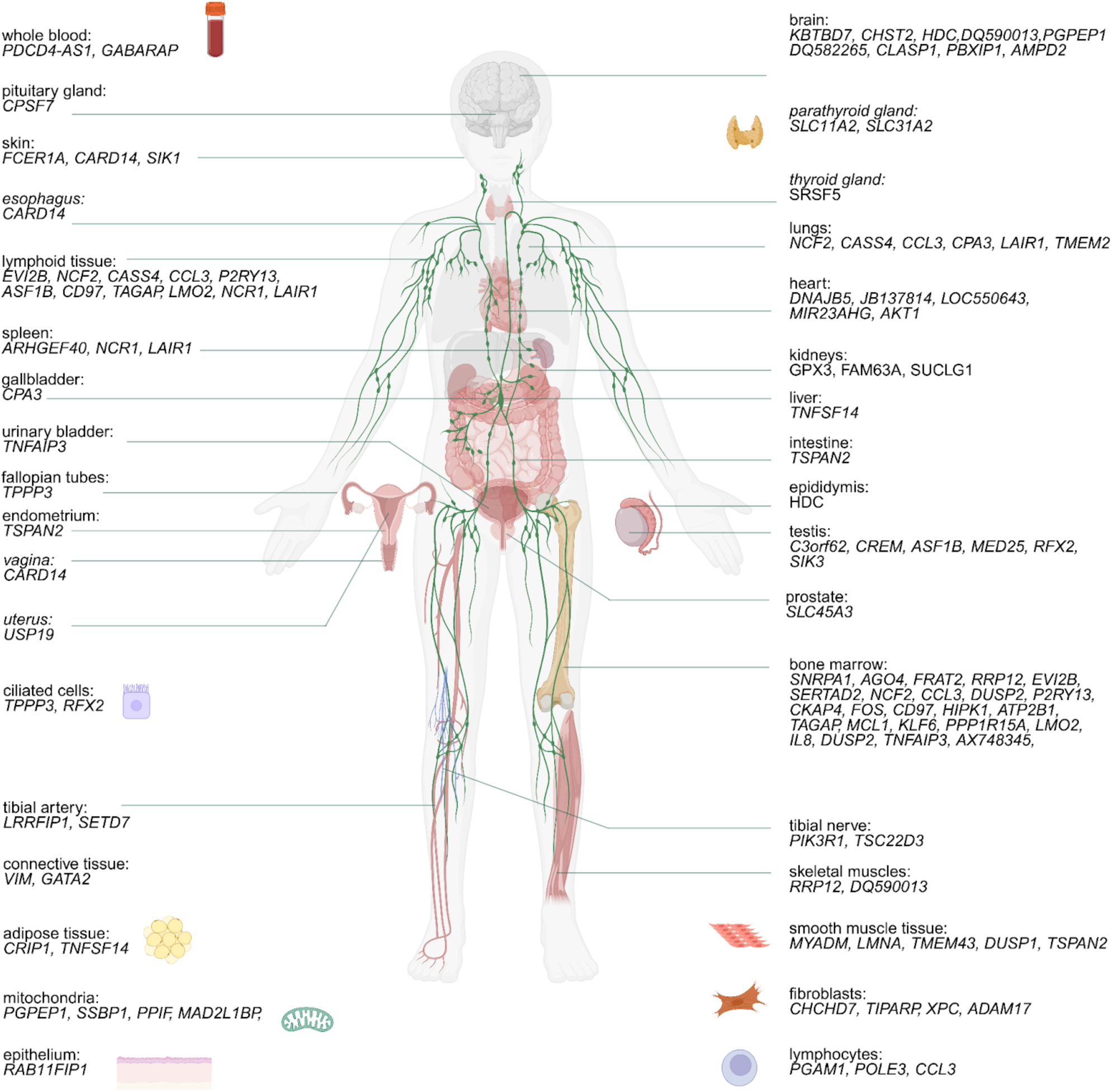
The tissues in which the 91 BP-DEGs are predominantly expressed.

### Gene Set Enrichment Analyses

After the differential expression analyses, we performed gene set enrichment analyses and analysed the resulting GO terms and pathways in which the DEGs were involved (see supplementary tables 5 and 6). Across all hypertension phenotypes, GO terms related to tube development, anatomical structure formation, and vascular development were consistently enriched, underscoring the critical role of blood vessel remodeling and angiogenesis in hypertension pathophysiology. Another recurrent theme for SBP, DBP and PP was GO terms for immune activation and inflammatory processes. GO terms unique to SBP included hydrogen peroxide catabolic process, hinting at the detection of oxidative stress. DBP terms overlapped significantly with SBP but included additional pathways related to signal transduction and cellular migration and adhesion. These terms highlight the importance of endothelial repair mechanisms and cell motility in maintaining vascular compliance. Terms related to gas transport, oxygen transport, and one-carbon compound transport were seen across the phenotypes emphasizing the critical role of metabolic and oxidative processes in hypertension. Additionally, we examined the KEGG pathways our genes of interest were involved in, and found similar results, i.e., pathways related to inflammation, blood vasculature and cell adhesion. However, for SBP two pathways were significant without direct links to hypertension, namely microRNAs in cancer and lysine degradation.

### Second Phase - Functional Evidence by Integrating Other Omics Layers

Once we found unique genes that were interesting to study further for hypertension, i.e., the 91 BP-DEGs, we were interested to find out if the expression of these genes was mediated by SNPs or CpG sites that were already reported for hypertension or its related phenotypes. Additionally, we assessed whether gene expression was causal for blood pressure changes or the other way around. Figure 3 provides a snapshot of the three analyses we performed

**Figure 3:**
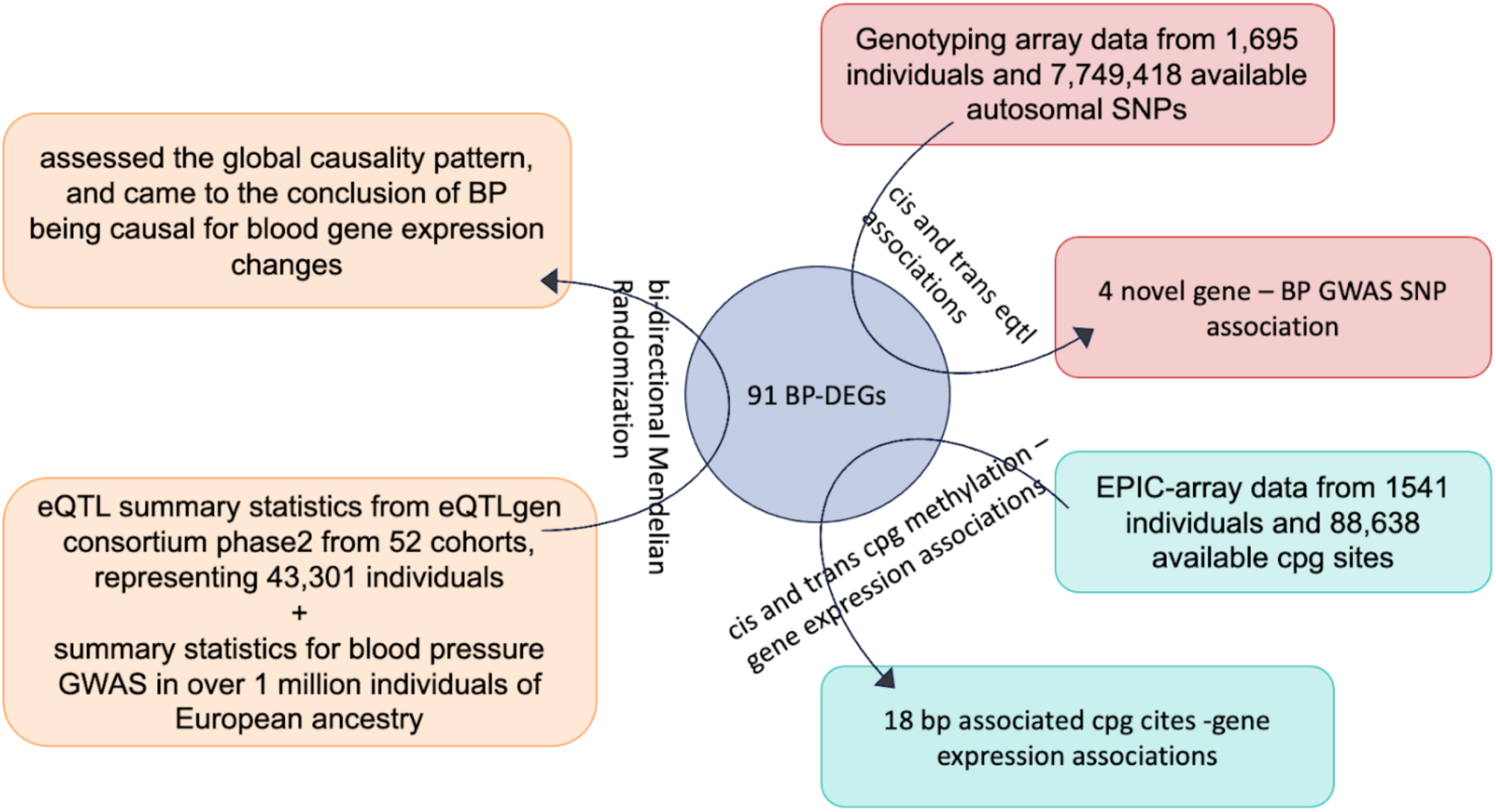
Overview of the analyses in phase 2.

### eQTL Analysis

We identified significant *cis* associations for 38 genes (see supplementary Table 7), and significant *trans* associations for 6 genes after Bonferroni correction (see supplementary Table 9). After finemapping, using linkage disequilibrium (LD) matrices computed from the dosages of the same genotyping array data, the number of *cis*-eQTLs was reduced to 1,852 (see supplementary Table 8), leaving only the SNPs that had in sum at least a 95% probability of including a causal variant modifying the expression of the respective gene. The number of *trans*-eQTLs was reduced to 106 (see supplementary Table 10). To put these findings in context, we performed a lookup of all these eQTLs in the GWAS catalogue and we found some associations with blood pressure traits along with some other cardiovascular traits, which could be observed in detail in supplementary tables 7 to 10. We found 11 *cis*-eQTLs for 5 genes and 3 *trans*-eQTLs for 2 genes in the GWAS catalogue associated with blood pressure indicating a causal relationship (see Table 3). The beta values for all the eQTLs are negative indicating that each additional copy of the alternative allele is correlated with a decrease in gene expression. Out of these 14 eQTLs, all were reported in GTEX except for 4, i.e., rs210131, rs9893005, rs35565381, and rs11959615, shown in red in the table.

**Table 3:** The cis/trans eQTLs that have also been reported in GWAS for hypertension or its related phenotypes. Underlined SNPs were also significant after LD fine-mapping. Novel hits are shown in red.

| eQTL | Variant- risk allele | BP-DEGs | p-value | beta | SE | Annotated gene | GWAS trait |
| --- | --- | --- | --- | --- | --- | --- | --- |
| *cis* | <u>rs6766740-A</u> | TMEM43 | 1.4E-11 | -0.12 | 0.02 | *TMEM43*, *RP11-434D12.1* | PP |
| *cis* | **rs210131** | **HSD17B8** | **5.5E-11** | **-0.27** | **0.04** | **<u>BAK1</u>** | **SBP** |
| *cis* | <u>rs111748634-C</u> | POLE3 | 6.4E-11 | -0.29 | 0.04 | *C9orf43* | SBP |
| *cis* | <u>rs111841972-A</u> | POLE3 | 6.7E-11 | -0.29 | 0.04 | *C9orf43* | SBP, PP |
| *cis* | rs111296842 | POLE3 | 5.2E-10 | -0.29 | 0.05 | *RGS3* | SBP |
| *cis* | rs12624154 | LRRFIP1 | 1.6E-09 | -0.11 | 0.02 | *LRRFIP1* | Hypertension |
| *cis* | rs9636202-A | PGPEP1 | 1.8E-09 | -0.17 | 0.03 | *PGPEP1* | Mean arterial pressure |
| *cis* | rs12459965-T | PGPEP1 | 3.5E-09 | -0.17 | 0.03 | *PGPEP1* | SBP, DBP, PP |
| *cis* | rs17724992-G | PGPEP1 | 6.2E-09 | -0.16 | 0.03 | *PGPEP1* | SBP |
| *cis* | rs34518929-A | PGPEP1 | 6.5E-09 | -0.16 | 0.03 | *PGPEP1* | SBP |
| *cis* | rs66978877-T | PGPEP1 | 7.1E-09 | -0.16 | 0.03 | *PGPEP1* | Hypertension |
| *trans* | **rs9893005-G** | **IL8** | **7.2E-13** | **-0.19** | **0.03** | ***PIGL*** | **SBP, DBP** |
| *trans* | **rs35565381-T** | **IL8** | **9.0E-13** | **-0.20** | **0.03** | ***PIGL*** | **DBP** |
| *trans* | **<u>rs11959615</u>** | **GPX3** | **2.8E-13** | **-0.20** | **0.03** | **<u>HTR4</u>, <u>ADRB2</u>** | **SBP** |

**Table 4:**
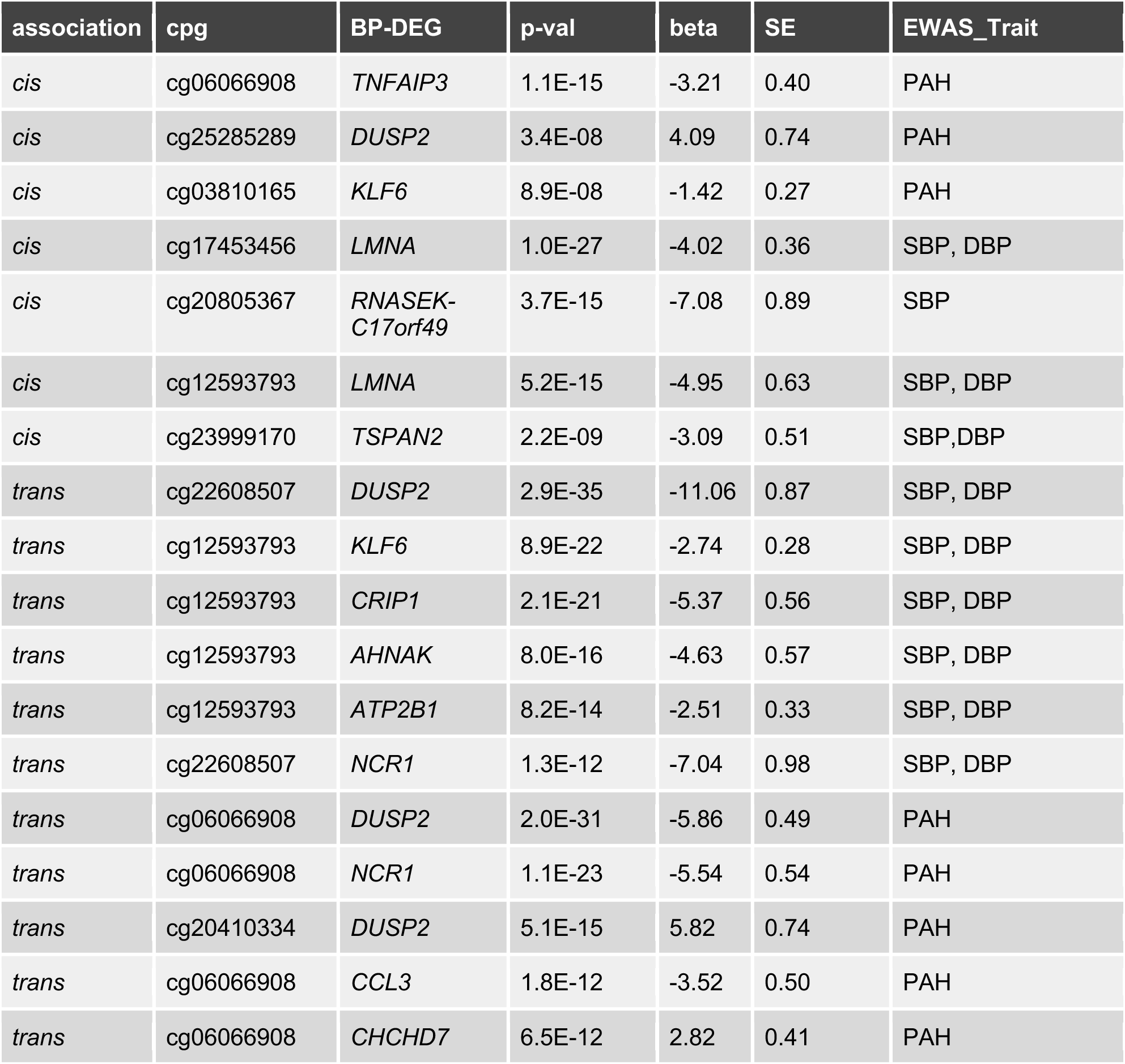
The cis and trans methylation-expression associations that have also been reported in EWAS for hypertension or its related phenotypes.

| association | cpg | BP-DEG | p-val | beta | SE | EWAS_Trait |
| --- | --- | --- | --- | --- | --- | --- |
| cis | cg06066908 | *TNFAIP3* | 1.1E-15 | -3.21 | 0.40 | PAH |
| cis | cg25285289 | *DUSP2* | 3.4E-08 | 4.09 | 0.74 | PAH |
| cis | cg03810165 | *KLF6* | 8.9E-08 | -1.42 | 0.27 | PAH |
| cis | cg17453456 | *LMNA* | 1.0E-27 | -4.02 | 0.36 | SBP, DBP |
| cis | cg20805367 | *RNASEK-C17orf49* | 3.7E-15 | -7.08 | 0.89 | SBP |
| cis | cg12593793 | *LMNA* | 5.2E-15 | -4.95 | 0.63 | SBP, DBP |
| cis | cg23999170 | *TSPAN2* | 2.2E-09 | -3.09 | 0.51 | SBP,DBP |
| trans | cg22608507 | *DUSP2* | 2.9E-35 | -11.06 | 0.87 | SBP, DBP |
| trans | cg12593793 | *KLF6* | 8.9E-22 | -2.74 | 0.28 | SBP, DBP |
| trans | cg12593793 | *CRIP1* | 2.1E-21 | -5.37 | 0.56 | SBP, DBP |
| trans | cg12593793 | *AHNAK* | 8.0E-16 | -4.63 | 0.57 | SBP, DBP |
| trans | cg12593793 | *ATP2B1* | 8.2E-14 | -2.51 | 0.33 | SBP, DBP |
| trans | cg22608507 | *NCR1* | 1.3E-12 | -7.04 | 0.98 | SBP, DBP |
| trans | cg06066908 | *DUSP2* | 2.0E-31 | -5.86 | 0.49 | PAH |
| trans | cg06066908 | *NCR1* | 1.1E-23 | -5.54 | 0.54 | PAH |
| trans | cg20410334 | *DUSP2* | 5.1E-15 | 5.82 | 0.74 | PAH |
| trans | cg06066908 | *CCL3* | 1.8E-12 | -3.52 | 0.50 | PAH |
| trans | cg06066908 | *CHCHD7* | 6.5E-12 | 2.82 | 0.41 | PAH |

### CpG Methylation-RNA Association

Since DNA methylation is known to influence gene expression, we integrated EPIC-array data from 1,541 participants and identified significant CpG sites linked to gene expression changes in BP-DEGs (see supplementary tables 11 to 13). We identified significant *cis* associations for 47 genes and *trans* associations for 68 genes. To investigate these CpG sites further, we performed a lookup in the EWAS Catalogue. We found 4 *cis* and 6 *trans* associations linked to blood pressure phenotypes and 3 *cis* and 5 *trans* associations related to pulmonary arterial hypertension (PAH), as shown in Table 3. It is worthwhile to note that the genes reported here to be significantly associated with the CpGs reported for essential hypertension traits in the EWAS Catalogue are already reported for hypertension in the literature, except for *RNASEK-C17orf49.* These results could explain how the methylation at the BP-linked CpG sites is responsible for BP changes, highlighting the importance of these genes for hypertension.

It is well established that DNA methylation is subject to change in response to environmental/lifestyle factors and can, in turn account for alterations in gene expression^26–28^. We explored which environmental/lifestyle factors were most frequently reported for all our significant *cis* and *trans* CpGs in the EWAS Catalogue. Smoking was the most commonly associated factor, followed by alcohol consumption.

### Mendelian Randomization

To assess whether genetically determined gene expression of the BP-DEGs is causal for the blood pressure phenotypes or vice versa, two-sample bidirectional MR was conducted. The detailed results including the number of instruments used in each analysis can be seen in supplementary tables 14 and 15.

According to the forward MR, when genetically determined gene expression was the exposure and blood pressure phenotypes were the outcomes, we found no significant results for SBP. For DBP as outcome, we found *PBXIP1, GABARAP, and AMPD2* to be significantly causal for DBP. For PP as outcome, we found *PBXIP1, NCF2, TMEM43, MIR23AHG* and *USP19* to be significantly causal for PP. For all the forward MR analyses, there was no significant directional pleiotropy or heterogeneity between the instruments. For reverse MR, when the exposure was blood pressure phenotypes and genetically predicted expression of BP-DEGs as outcomes, SBP was found to be significantly causal for the expression of 22 genes (see Table 5). DBP was found to be significantly causal for 10 genes and PP was found to be significantly causal for 8 genes*. KLF6* and *TSPAN2* were the only 2 genes significant for all the blood pressure phenotypes for reverse MR. For all these reverse MR analyses, there was no significant directional pleiotropy detected; however, there was heterogeneity between the instruments, which could be because of the large number of instruments. The genes *PBXIP1* and *MIR23AHG* exhibit bidirectionality. *PBXIP1* is significant in forward MR for both DBP and PP and also in the reverse MR for both SBP and DBP. *MIR23AHG* is significant for forward MR for PP and also for reverse MR for both SBP and PP. For more details on the number of instruments, p-values, pleiotropy and heterogeneity variables, see supplementary tables 14 and 15.

**Table 5:**
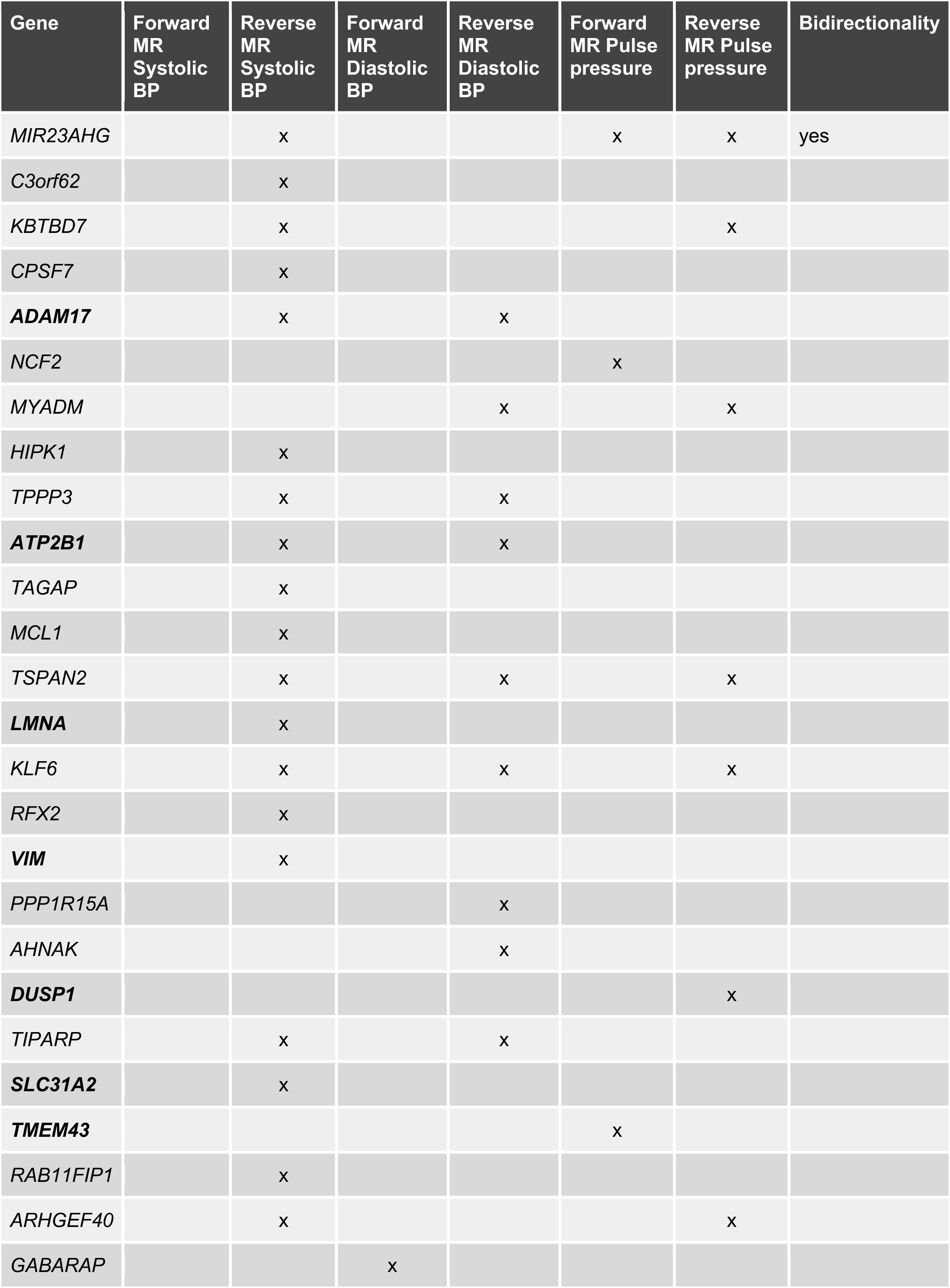
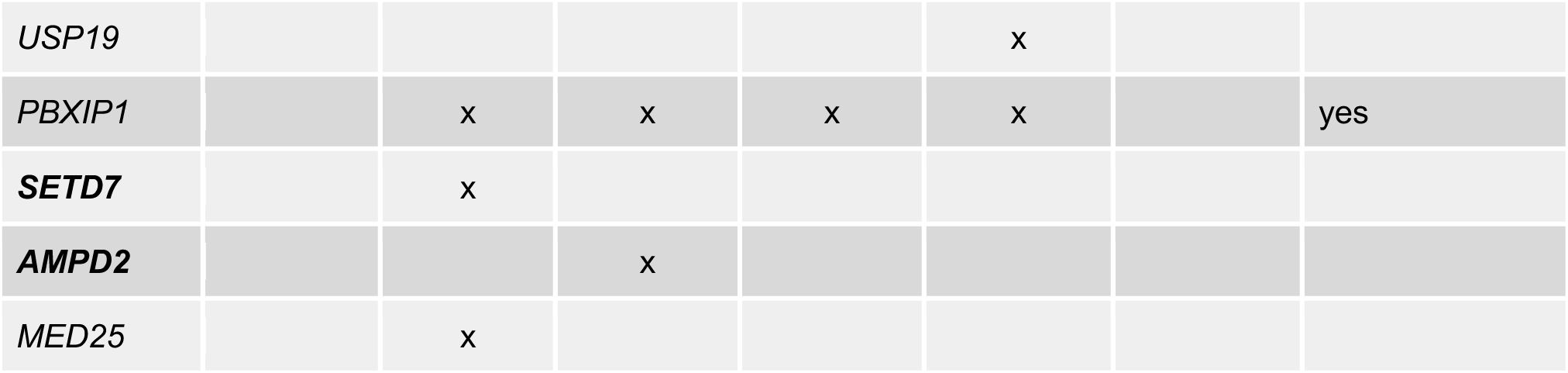
Overview of results from the Mendelian randomization analysis. x marks the significant genes after multiple testing in the respective analyses. The bold genes are part of the druggable genome.

We performed Fisher’s exact test to assess differences in the number of causal associations between forward and reverse MR analyses for the three BP traits. The results showed a highly significant difference for SBP (p = 2.015 × 10⁻⁶), suggesting a strong unidirectional effect where SBP is causal for gene expression rather than the other way around. For DBP, there was a non-significant trend (p = 0.081), and PP also showed no significant difference (p = 0.549). These results suggest that BP is causal for gene expression. Our findings are in line with existing literature^29^.

## Discussion

In the presented study, we analysed RNA-seq data from the KORA FF4 cohort in two phases. Phase one focused on gene discovery and literature review to determine what the genes are known for, which pathways they are involved in and which tissues exhibit the highest expression of the found genes. Phase two focused on gathering functional evidence by linking known BP-related genetic and epigenetic variations to gene expression. In phase two, we additionally assessed causality patterns between BP and gene expression, and in light of these findings proposed new, viable drug targets. In the subsequent paragraphs, we will highlight the most noteworthy findings from these analyses.

In the first phase, we came up with a list of 91 BP-DEGs. Out of the novel genes, we only focus on 7 genes that were also significant in the sensitivity analysis i.e., *CCT6P1, RNASEK-C17orf49, PDCD4-AS1, SNRPA1, AGO4, PGPEP1* and *MIR23AHG.* Out of these 7 genes, *MIR23AHG* (also known as *MIR23AHG*) shows the most promise for a functional follow-up to get insight into potential new blood pressure pathways, as it was significantly upregulated with the increase in both SBP and DBP in the sensitivity analysis and was the only novel gene significant for all three HTN phenotypes i.e. SBP, DBP and PP in the initial gene expression analysis. *MIR23AHG* is a long noncoding RNA (lncRNA) expressed the most in the heart (specifically in the aorta), which is a very relevant organ for hypertension and we can detect its presence in the blood. *MIR23AHG* is also significant in reverse MR for SBP and PP and forward MR for PP. This may imply a more complex directional causality, however, we have stronger statistical evidence in reverse MR because IVW method offers more power than a single-SNP Wald ratio test in forward MR. Additionally, we arrive at these results with Steiger filtering and no horizontal pleiotropy, making the reverse association robust. The increase in blood pressure also causes the gene expression of *MIR23AHG* to increase, advocating its biological importance. The genes *CCT6P1, PDCD4-AS1, SNRPA1* and *AGO4* are mostly reported in literature for cancers; further functional studies are required to discern their utility in hypertension and cardiovascular diseases.

When looking at the highest expression of the rest of the BP-DEGs in various tissues, we can highlight the systemic effects of blood pressure. The DEGs detected in whole blood might originate from circulating immune cells or other cell types that display the strongest expression in brain, lymphoid organs, bone marrow, blood vessels, muscle cells, kidneys, etc. Hypertension is a complex condition involving multiple organ systems, and looking at the different tissues highlights this even further.

The results of gene set enrichment analyses highlight processes such as vascular remodelling, which are consistent with known mechanisms underlying vascular stiffness and intimal thickening in chronic hypertension^30^. Due to endothelial damage in hypertension, inflammatory response in the vessels is well documented; our findings support the growing body of evidence linking inflammation and hypertension^31,32^. Pathways concerning oxidative stress are also reported in literature^33,34^. The KEGG pathways in which our genes of interest were dysregulated,such as those related to inflammation, blood vasculature and cell adhesion, have also been reported in literature^31,32,35^.

The eQTL and CpG methylation-gene expression analyses were meant to decipher if the SNPs and CpGs already implicated in hypertension have an influence on the gene expression of our 91 BP-DEGs already prioritized to play a role in hypertension. The result section shows the correlation of these SNPs and CpGs, highlighting some new findings. We would like to mention two main genes here, *PGPEP1* and *RNASEK-C17orf49* from our list of 7 novel genes mentioned above. The *PGPEP1* gene’s locus is known to be associated with growth differentiation factor-15 (GDF-15) concentration in the blood^36^. Animal studies indicate that GDF-15 exhibits anti-inflammatory properties and helps prevent the development of atherosclerosis^37,38^. In our study, *PGPEP1* is downregulated with the increase in pulse pressure and there are 5 blood pressure GWAS hits that also act as eQTLs for this gene; with the presence of the risk allele, the expression decreases (except for rs66978877, where the expression is higher when the risk allele is present). Here we can conclude that the risk allele genotypes may cause the expression of *PGPEP1* to decrease, and this combined with the fact that the expression of *PGPEP1* gets lower with pulse pressure, is potentially hindering the protective mechanisms in place to prevent the deleterious effects of high blood pressure on the body. Our MR analyses do not show a significant result for *PGPEP1*, and the possible explanation for this could be that the highest expression of this gene is detected in the brain according to GTEX, making whole blood a weaker tissue to consider when choosing instruments for gene expression for the MR analysis. Similarly, the gene *RNASEK-C17orf49* has a *cis* CpG site, reported in the EWAS Catalogue to be associated with blood pressure, where the increase in the degree of methylation is linked to decreased SBP and we see the same trend in the expression of *RNASEK-C17orf49* with diastolic blood pressure, implying an epigenetic mediation of gene expression and its potential involvement in hypertension.

When recommending potential therapy targets for hypertension or related comorbidities, we looked at the genes that were significant either in the forward MR or the reverse MR, in addition to being listed as druggable in the DGIdb. Only 9 genes fulfilled the criteria i.e., *TMEM43* and *AMPD2* for which gene expression causes changes in BP, and *ADAM17, ATP2B1, LMNA, VIM, DUSP1, SLC31A2* and *SETD7* for which BP is causal for changes in expression. While all these genes could be considered for therapy as we explain below, we would like to give priority to *LMNA* as it is the only gene that was significant in our sensitivity analysis for blood pressure. *LMNA* is significantly upregulated for BP in our differential expression analyses, and according to the MR analysis, high BP causes the expression of *LMNA* gene to increase. According to a recently published study, an increased amount of Lamin A (a gene product of *LMNA*) is associated with significantly impaired cardiac function, enlarged heart chamber of the left ventricle and reduced left ventricle wall thickness coupled with interstitial fibrosis^39^. Exploring *LMNA* as a potential drug target can help mitigate the adverse effects that come with hypertension. *ADAM17* has already been vetted for a therapy target for hypertension^40^. The *ATP2B1* gene locus has been heavily implicated in hypertension and has already been suggested as a drug target^41,42^. According to the forward MR results, an increase in expression of the *TMEM43* gene causes a decrease in PP, which makes it a promising target for therapeutic intervention, as a higher value of PP is associated with arterial stiffness and CVDs like atherosclerosis^43^. *TMEM43* has already been suggested as a potential drug target for arrhythmogenic cardiomyopathy type 5, where its high expression can delay the onset of the condition, but it has never been suggested for hypertension^44^. Increased expression of *AMPD2* causes an increase in DBP in our forward MR analysis and inhibiting the expression may be a good therapeutic approach for managing hypertension. However, in our differential expression analysis, *AMPD2* was downregulated only for men in the hypertension group, putting its drug target candidacy in question. *DUSP1* is listed as a drug target for an antidiuretic in the DGIdb, which helps to raise BP. In our differential expression analyses, *DUSP1* is upregulated in all the HTN phenotypes, suggesting the body’s protective responses in action to mitigate the harmful effects of hypertension. This is in line with the existing literature that expression of *DUSP1* has protective effects against cardiac ischemia^45^. An Increase in SBP causes an increase in the expression of *SLC31A2* according to our reverse MR results. It is upregulated for SBP and PP, and according to literature, it is also upregulated for atherosclerosis^46^. Since members of the solute carrier family such as the *SLC12A3,* are already drug targets for diuretics, incorporating *SLC31A2* in therapy could help slow down the adverse effects of hypertension and manage it well^47^. Following the same logic, an increase in SBP causes an increase in the expression of *VIM*, and increased expression of *VIM* has been associated with increased inflammation and coronary artery disease^48,49^. In our MR analysis, an increase in SBP causes an increase in the expression of *SETD7.* In literature, downregulation of *SETD7* reduces myocardial injury, infarct size, and mitochondrial ROS production, hence, it has already been proposed as a drug target for cardiovascular diseases and inflammation^50,51^. Repurposing/studying these genes as drug targets for hypertension could prove to be beneficial.

Our study has several strengths, i.e., a large sample size and the use of RNA sequencing data, which together enhance our power to discover novel genes. Increasing the sample size further could continue to improve the detection of these genes. Additionally, we leveraged multi-omics data collected from the same cohort, allowing us to integrate genetic, epigenetic, and transcriptomic layers. This multi-dimensional approach provides a more comprehensive understanding of how genetic variation and DNA methylation may mediate gene expression and ultimately contribute to hypertension. Future directions include the use of RNA-seq data from hypertension-relevant tissues, such as arteries, which may yield more tissue-specific insights. Incorporating longitudinal data could also help evaluate the predictive value of the candidate genes we found in the early detection of hypertension. For instance, developing an RNA-based risk score that aggregates expression profiles of novel and known genes in an independent cohort could enable early identification of high-risk individuals and inform preventative interventions. Overall, our findings highlight a set of promising candidate genes for further functional studies and represent a meaningful step toward better understanding and controlling hypertension.

## Methods

### Study Population

The conducted study utilized data from the KORA FF4 study, a population-based survey conducted in Augsburg, Southern Germany, in 2013/2014. As the second follow-up to the KORA S4 health survey (1999-2001)^52^, 2,279 of the original 4,261 participants took part. At the study center, participants filled self-administered questionnaires, took part in computer-assisted interviews with trained nurses, and underwent a standardized physical examination that included anthropometric measurements, oral glucose tolerance test (OGTT) and blood pressure measurements. Blood pressure measurements were recorded using a HEM-705CP device with 2 cuff sizes (Omron Corporation, Tokyo, Japan). After resting for at least 5 min, three measurements with 3 min rest in between were recorded. The average of the 2nd and 3rd measurements were used for analysis. Participants were asked to bring in their medication and usage was recorded for the past 7 days. Blood samples were also collected that were later used for different omic measurements. Out of the 2,279 blood samples only 2,013 samples with the best RIN (RNA integrity number) and concentration were used. After rigorous QC only 1,796 participants qualified for the study, the characteristics of whom can be seen in Table.1. The study was approved by the Bavarian Chamber of Physicians’ Ethics Committee, and all participants that are included in the study provided written informed consent.

### Quality Control of the Omics Data Transcriptomics Data

After RNA isolation using PAXgene® Blood RNA Kit, RNA integrity number (RIN) was measured using the Agilent 2100 Bioanalyzer system. RNA samples with RIN values of approximately 6 or more were selected for mRNA sequencing (poly-A selected). The libraries were prepared using the Illumina stranded mRNA prep ligation kit (Illumina), following the kit’s instructions. After a final QC, the libraries were sequenced in a paired-end mode (2×100 bases) in the Novaseq6000 sequencer (Illumina) with a depth of ≥ 40 Million reads per sample. After demultiplexing, FASTQ files from each sample were processed using standard tools^53^. Alignment to UCSC Genome Browser hg19 human reference genome using STAR v2.4.2a^54^. Unaligned reads are discarded. Sequencing QC was done using RNASeQC v1.1.8.1^55^. Properly aligned reads are then processed with HTSeq-count v0.6.1^56^ to generate read counts which can be interpreted as quantified gene expression. The reads are then normalized for exon length and total sequencing yield to generate Fragments Per Kilobase of transcript per Million mapped reads (FPKM), and this is done through dividing the fragments per gene by the product of length of the gene in kilobase and million reads sequenced.

After sequencing QC, samples QC was done. Samples with < 30 million reads were discarded. Exonic, intronic, intragenic, intergenic and rRNA rates calculated by RNASeQC were examined for outliers but no such outliers were found and no samples were excluded based on these. Only the genes with FPKM of ≥ 1 in at least 5% of the samples were selected. Number of the selected genes in each sample was calculated. Samples having less than 5,750 genes (determined by visual inspection of a histogram of gene count distributions) were excluded. Sex mismatches in the phenotype tables and those discerned from looking at the expression of XIST and UTY genes were also excluded.

### Genotyping Array Data

Genome-wide SNP data were obtained using an Affymetrix Axiom array. NCBI build 37 was used for calling the SNPs. Samples with high levels of missing SNPs i.e individuals not having genotyping calls for at least 97% of the SNPs are removed. Gender checks were made to correct for sample swaps by observing mismatches of phenotypic and genetic gender. These samples were removed. Samples with high or low heterozygosity rates were excluded (5*standard deviation of mean heterozygosity rate). SNPs with low genotyping calls, i.e. SNPs not present in at least 97% of the samples were removed. SNPs which deviate from Hardy–Weinberg equilibrium were excluded (p-value<5×10^−10^). SNPs are imputed using the Michigan Imputation Server and software minimac3. As a Reference panel, Haplotype Reference Consortium is used. To correct for multiple testing and to select only SNPs with high statistical power in the analysis, only SNPs with a minor allele frequency above 1% are considered.

### Methylation Data

Genomic DNA (750 ng) was bisulfite converted using the EZ-96 DNA Methylation Kit. Subsequent methylation analysis was performed on an Illumina iScan platform using the Infinium MethylationEPIC BeadChip according to standard protocols provided by Illumina. GenomeStudio software version 2011.1 with Methylation Module version 1.9.0 was used for initial quality control of assay performance and for generation of methylation data export files. Further quality control and preprocessing of the data were performed in R v3.5.1, with the package minfi v1.28.3^57^ and following primarily the CPACOR pipeline^58^. Raw intensities were read into R and background corrected. Probes with detection p-values

>0.01 were set to missing. Samples with mismatched reported sex and that estimated from the methylation data were excluded. Samples with median intensity <50% of the experiment-wide mean, or <2000 arbitrary units were removed. Samples with >5% missing values on the autosomes were removed. A total of 59631 probes i.e probes with SNPs with minor allele frequency >5% at the CG position or the single base extension as given by minfi; and 5786 with >5% missing values. A total of 806228 probes were kept for downstream analyses. Quantile normalization (QN) was then performed separately on the signal intensities divided into the 6 probe types: type II red, type II green, type I green unmethylated, type I green methylated, type I red unmethylated, type I red methylated^57^. The transformed intensities were then used to generate methylation beta values, a measure from 0 to 1 indicating the percentage of cells methylated at a given locus.

### Differential Expression Analyses

Differential expression analysis was performed using voom to transform raw RNA-seq read counts to log counts per million (log-cpm) with associated precision weights, followed by linear modeling and empirical Bayes procedure using limma^59^. We used the following linear regression model for SBP, DBP and PP:

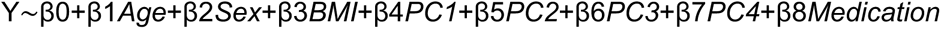

where *Y* is the gene expression, *Age* denotes the age of the subjects in years, *BMI* denotes the body mass index, *Sex* denotes the reported sex and *Medication* denotes whether the subjects were on any antihypertensive medication. Additionally, we decided to add the first four principal components(PCs) from PCA analysis done using DEseq2 package in RStudio on normalized counts to correct for technical covariation. We chose the top four principal components explaining most of the variance. When plotting the PCs it was clear that PC2 correlates to the percentage of white blood cells (lymphocytes and neutrophils) and PC3 was batch effect. PC1 and PC4 could not be discerned even after testing a multitude of biological covariates and technical covariates such as RIN scores and concentration of RNA used. When testing for HTN, we excluded the intake of antihypertensive medication variable from the regression model because the use of medication was already included in the definition of HTN. When doing the age specific analyses we took out the sex variable from the regression model. The p values produced from the regression models were adjusted for multiple testing using the Bonferroni method. Genes with Padj < 0.05 were considered differentially expressed.

### Pathway Analysis

We performed Gene Set Enrichment Analysis (GSEA) by querying Gene Ontology (GO) terms using ranked Gene Set on fold changes using RStudio function gseGO from the clusterprofiler package^60^. GO term assignments were available in the Molecular Signatures Database (MSigDB)^61^. For the biological pathways, we also looked at Kyoto Encyclopedia of Genes and Genomes (KEGG) by using the gseKEGG function from the clusterprofiler package.

### eQTL and CpG Methylation-Gene Expression Anaylses

To decipher the expression changes taking the genetic makeup into account, we integrated genotyping array data from 1,695 individuals included in our differential expression studies. The total number of imputed autosomal SNPs available to us was 7,749,418 and we saw the association of these with our 91 BP-DEGs. For the *cis*-eQTLs analysis we performed 489,408 tests, hence Bonferroni corrected significance threshold was < 0.01/489,408 (<2.04×10-8). For the *trans*-eqtls analysis we performed 689,208,794 tests, hence Bonferroni corrected significance threshold was < 0.01/689,208,794 (<1.5×10-11). For the CpG methylation-gene expression analysis we integrated EPIC-array data from 1541 individuals and 88,638 available CpG sites, and saw the association of the degree of methylation at the CpG sites and expression changes in our 91 BP-DEGs. For the *cis* associations we performed 88,638 tests, hence Bonferroni corrected significance threshold was < 0.01/88,638 (<1.1×10-7). For *trans* associations, we performed 65,642,223 tests, hence Bonferroni corrected significance threshold was < 0.01/65,642,223 (<1.5×10-10).

For both eQTL and mQTL calculations we used an R package called MatrixeQTL v2.3^62^. For eQTL calculation, the FPKMs underwent quantile normalization to align the distributions of the samples and gene-wise quantile normalization. In this last step, values are aligned per gene to the quantiles of the normal distribution, which is done to be able to use linear models for the QTL analysis. Following the normalizations, to minimizing the impact of measured and unmeasured confounders, we adjust the normalized counts by calculation residuals of the linear models of expression values corrected by age, sex, bmi, antihypertensive medication, and PCs 1 to 4, just like we did for our differential expression models. We did this because the genotypes are unaffected by the confounders we use for the expression data^63^. We used these residuals and dosages of the SNPs from the QCed genotyping array data to calculate the *cis* and *trans* associations with our 91 BP-DFGs. The p-values were corrected for multiple testing by Bonferroni method i.e. p value less than 0.01/number of tests performed was taken as significant. For mQTL calculation the same approach was used, when calculating the residuals first 10 PCs and percentage of neutrophils, basophiles, eosinophils and monocytes were used as recommended in the literature. For expression data the normalized counts were used to calculate residuals by using only the first 4 PCs explaining the most variance, and since we were comparing the same subjects, effects of age, sex and BMI were corrected when calculating the association of gene expression and methylation.

### LD Fine-mapping for eQTLs

For each of our genes, Bonferroni-significant eQTL SNPs (p < 0.01) were searched for in the genotyping array data to make a matrix of dosages, this information was later used to build a correlation matrix. The correlation matrix was then mean-centered and scaled to unit variance to be used to correct for LD. We also incorporated gene expression matrix, with all the technical and biological variation corrected for, and ran a univariate linear regression to compute effect sizes and standard errors for each SNP. We used susie_rss function from the susieR package to calculate the posterior inclusion probabilities (PIPs) and credible sets by using the default settings. SuSie(Sum of Single Effects) is a statistical method used to fine-map complex traits by identifying multiple potentially causal variants from a set of genetic variants. It uses a Bayesian framework to estimate the posterior distributions of effect sizes for each variant, incorporating information on linkage disequilibrium. It calculates PIPs for each variant, which reflect the likelihood that a given variant is truly causal^64,65^. Each variant is assigned a credible set, the larger the size of a set, the less sure we are about the probability of variants being causal. This explains the low PIPs for eQTLs in bigger credible sets.

### MR Analysis

Causality was assessed in BP-DEGs. The following 4 methods were tested, i.e MR Egger (ME), weighted median (WMe), inverse variance weighted (IVW), and Wald test (WT). We deemed a result significant if multiple testing corrected p-value of IVW was significant, and the direction of effect was consistent in IVW, ME and WMe. In the case of just one available instrument, significant p-value after multiple testing correction of WT was taken as significant. We tested 70 genes for SBP, 42 genes for DBP and 32 genes for PP, that made the significance thresholds for SBP <0.05/70 (<7.1e-04), for DBP < 0.05/42 (<1.2e-03) and for PP <0.05/32 (<1.6e-03). These number of genes per blood pressure phenotype come from our differential expression analysis of the whole cohort, sex specific differential expression analyses and if a gene was differentially expressed for HTN, we added it for both SBP and DBP. All MR analyses were conducted using MRBase as implemented in the R packages TwoSampleMR v.4.22^66^.

For the forward MR, gene expression was the exposure and blood pressure was the outcome. Instruments for gene expression were taken from summary statistics from eQTLgen consortium phase2 where genome-wide eQTL analysis was performed in 52 cohorts, representing 43,301 individuals (see supplementary file). Per gene, only the significant *cis*-eQTL SNPs with genome wide significance (5e-8) were chosen. To reduce pleiotropy potentially violating the MR assumptions, all the SNPs reported to have genome wide significance in a GWAS meta-analysis of BMI in ∼700000 individuals of European ancestry were excluded^67^. We additionally excluded the SNPs reported for BMI in the GWAS catalogue. This list of SNPs was searched for in the summary statistics for blood pressure in over 1 million individuals of European ancestry^10^. After shortlisting the common SNPs, LD clumping was performed with r2 of 0.01 and a genome window of 10,000 kbps. We also excluded SNPs left after clumping if their proxies were reported for BMI in the GWAS catalog using the LDtrait online tool from NIH^68^. We defined proxies using an R² threshold of 0.8 within a ±500,000 base pair window. The chosen set of SNPs were then harmonized i.e SNPs with alleles on incompatible strands as assessed by looking at alternate allele frequencies, and Palindromic SNPs with minor allele frequency close to 50% were excluded before the MR calculation. For the reverse MR, we basically used the same steps, the only difference was that for instruments we used genome-wide significant SNPs from the summary statistics for blood pressure in over 1 million individuals of European ancestry^10^.

## Associated Data

### Supplementary Materials

Supplementary file: Word File comprising of supplementary information referred to in the main text (36 kb).

Supplementary Tables: Excel file comprising of 15 tables giving detailed information on the results summarized in the manuscript (XLSX 24,324 kb).

### Author Contributions

Conceptualization, A.Farzeen, H.Prokisch, C.Gieger; RNA extraction, N.Lindemann; Upstream QC of RNA-seq data, R.Berutti, M.Pavlov; methodology, A.Farzeen; software, A.Farzeen; validation, A.Farzeen, C.Gieger; formal analysis, A.Farzeen; investigation, A.Farzeen; resources, C.Gieger, M. Waldenberger, B.Linkohr; data curation, A.Farzeen; writing— original draft preparation, A.Farzeen; writing—review and editing, A.Farzeen, H.Prokisch, C.Gieger, A.Teumer; visualization, A.Farzeen, S.Sharma, M.Fuchs; supervision, A.Peters, S.Sharma, H.Grallert, J.Winkelmann, L.Mägdefessel, A.Teumer, H.Prokisch, C.Gieger; project administration, B.Linkohr, H.Prokisch, C.Gieger; funding acquisition, H.Prokisch, C.Gieger, B.Linkohr, H.Schunkert, M.Scheidt, A.Peters. All authors have read and agreed to the published version of the manuscript.

### Funding

The KORA study was initiated and financed by the Helmholtz Zentrum München – German Research Center for Environmental Health, which is funded by the German Federal Ministry Education and Research (BMBF) and by the State of Bavaria. Data collection in the KORA study is done in cooperation with the University Hospital of Augsburg. This study was funded by the Bavarian State Ministry of Health, Care and Prevention through the research project DigiMed Bayern (www.digimed-bayern.de). A.Teumer has been funded by the Deutsche Forschungsgemeinschaft (DFG, German Research Foundation) – 542489987

### Institutional Review Board Statement

The study was conducted according to the guidelines of the Declaration of Helsinki and approved by the KORA Board (PV K022/23o).

### Informed Consent Statement

Written informed consent has been obtained from the patients to publish this paper.

### Data Availability Statement

The KORA and the RNA sequence datasets are not publicly available but can be accessed upon application through the KORA-PASST use and access hub subject to KORA Board approval (https://helmholtz-muenchen.managed-otrs.com/external/).

## Acknowledgments

We thank all participants for their long-term commitment to the KORA study, the staff for data collection and research data management and the members of the KORA Study Group (https://www.helmholtz-munich.de/en/epi/cohort/kora) who are responsible for the design and conduct of the study. Library preparation and sequencing was performed at the Helmholtz Zentrum München (HMGU) by the Genomics Core Facility. Methylation analysis and initial quality control was performed at the Genome Analysis Center (GAC), Helmholtz Zentrum München. Further quality control and preprocessing of the methylation data was performed by Thomas Delerue from the Department of Molecular Epidemiology, Institute of Epidemiology, Helmholtz Zentrum München. We also thank the eQTLGen Consortium for providing the summary stats for MR analyses.

