## Supplementary Information for "Novel and Known Genetic Players in Hypertension: From Gene Expression to Striking Insights"

#### Table of Contents:

Literature Overview for the Genes of Interest (91 BP-DEGs) 3
Methods Used to Generate eQTLGenConsortium Phase 2 Summary Statistics 5
Author Information For eQTLGen Consortium 7
References: 14

### Literature Overview for the Genes of Interest (91 BP-DEGs)

Elaborating on the overview from table 2, of our 91 BP-DEGs, 30 gene have been previously reported for hypertension or its phenotypes in previously published gene expression studies, the direction of effects was largely consistent except for *SLC45A3, IL8* and *TNFSF14* because in our study they're all downregulated for increasing blood pressure as opposed to the literature[1–7]. 10 genes are reported for hypertension in mouse models[8–16]. *AKT1* has been shown to have a protective effect for hypertension, consistent with our results as well[8]. *DUSP2* is reported to be upregulated in chronic hypertension in rats, in addition to being involved in human heart failure[9]. *TNFAIP3* is linked to pulmonary hypertension in mice, however the direction seems to be inconsistent with our study as the knockout mice of *TNFAIP3* develop pulmonary hypertension[10]. The direction also seems to be inconsistent in mice knockout studies for *PPIF, SUCLG1, SRSF5, SIK1* and *SIK3*[11,14–16]. Proteins of the genes *PGAM1, ADAM17, CASS4* and *CCL3* are reported for hypertension[17–19]. Although *CASS4* is majorly known for Alzheimer's disease and cancer, it is heavily implied that it may have a part to play in hypertension by proxy of its paralogue *NEDD9*[18]. SNPs in *CARD14, SLC11A2, CHCHD7, SERTAD2, PIK3R1* and *NCF2* have been linked to hypertension[20–25].

Two genes from our BP-DGEs are reported for preeclampsia, *LAIR1* is downregulated in preeclampsia which is also consistent with our results, and polymorphisms in the gene *XPC* have also been linked to preeclampsia[26,27]. *MAD2L1BP* and *GATA2* have been reported to be downregulated in pulmonary arterial hypertension, consistent with our results[28,29]. As cardiovascular diseases and hypertension are closely linked, we expected to find an overlap between our BP-DEGs and genes linked to some cardiovascular phenotypes. For example, the gene *TMEM2* was reported to be elevated in ST-elevation myocardial infarction (STEMI)[30], *CREM* is linked to cardiomyocyte hypertrophy, fibrosis, and left ventricular dysfunction in mice[31], *ASF1B* may be indirectly involved in myocardial infarction[32], *LRRFIP1* in thrombosis formation, inflammation, obesity and cancer[33]. *PBXIP1* is a marker for arterial fibrillation and CAD[34]. *HDC* is known for chronic heart failure[35], *MED25* is known for heart malformation and lipid metabolism and *POLE3* for lower extremity artery disease[36,37]. *P2RY13* may contribute to the inhibition of cardiac sympathetic drive, hence making it suitable for therapies aimed at treating cardiovascular conditions like stroke and myocardial infarctions[38]. In our study *P2RY13* is downregulated with increased blood pressure consistent with its reported function. *USP19* acts as a negative regulator of pathological cardiac hypertrophy, however, in our study it is upregulated[39]. The deficiency of *GPX3* is linked to cardiovascular diseases such as carotid atherosclerosis, the direction is consistent with our findings[40]. *SETD7* is upregulated in cardiometabolic heart failure with preserved ejection fraction, and it's also upregulated for blood pressure in our study[41]. *CPA3* is upregulated in patients with carotid atheroma[42]. *SSBP1* is upregulated in Keshan disease which is a kind of endemic cardiomyopathy, but we sound it to be downregulated for blood pressure in our analysis[43]. *CKAP4* is a marker for fibroblast activation in the heart that in turn causes ischemic injury, and vascular calcification in kidney disease[44,45].

21 have never been reported for hypertension or any cardiovascular diseases. Literature search did reveal, however, that *CLASP1* is involved in microtubule stabilization in cells[46], one could imply its function in arterial stiffness paramount to increasing blood pressure. *SNRPA1* is involved in renal carcinoma and inflammation[47]. *C3orf62*, *LOC550643*, *DQ590013* and *EVI2B* are associated with cancers[48–51]. Additionally, *LOC550643* is a non-coding RNA that's involved in mRNA turnover and nonsense-mediated decay and in our study, it is downregulated with increasing blood pressure. Variants in the gene *FCER1A* are reported for atopic eczema and asthma, and *CHST2* for rheumatoid arthritis[52,53]. *AX748345* is an antisense to *SPNS3* which is patented for PAH detection. All these genes represent positive avenues for blood pressure research.

### Methods Used to Generate eQTLGenConsortium Phase 2 Summary Statistics

All 52 cohorts performed cohort-specific analyses as outlined in the eQTLGen analysis cookbook (https://eqtlgen.github.io/eqtlgen-web-site/eQTLGen-p2-cookbook.html). Genotype quality control was performed according to standard bioinformatics practices and included quality metric-based variant and sample filtering, removing related samples, ethnic outliers and population outliers. Genotype data was converted to genome build hg38 if not done so already and the autosomes were imputed using the 1000G 30x WGS reference panel (64) (all ancestries) using the eQTLGen imputation pipeline (https://github.com/eQTLGen/eQTLGenImpute).

Like the genotype data, gene expression data was processed using the eQTLGen data QC pipeline (https://github.com/eQTLGen/DataQC). For array-based datasets, we used the results from the empirical probe mapping approach from our previous study6 to connect the most suitable probe to each gene which has previously been to show expression in the combined BIOS whole blood expression dataset. Per microarray platform, genes that showed negative correlation in the empirical probe mapping approach were excluded. Raw expression data was further normalized in accordance with the expression platform used (quantile normalization for Illumina expression arrays and TMM95 for RNA-seq) and inverse normal transformation was performed. Gene expression outlier samples were removed and gene summary information was collected for filtering at the central site. Samples for whom there were mismatches in genetically inferred sex, reported sex, or the expression of genes encoded from sex chromosomes were removed. Similarly, samples with unclear sex, based on genetics or gene expression were removed.

An adaptation of the HASE framework96 was used to perform genome-wide meta-analysis. For genome-wide eQTLs analysis, this limits the data transfer size while ensuring participant privacy. At each of the cohorts, the quality controlled and imputed data was processed and encoded so that the individual level data can no longer be extracted, but while still allowing effect sizes to be calculated for the linear relationship between variants and gene expression. (https://github.com/eQTLGen/ConvertVcf2Hdf5 and https://github.com/eQTLGen/PerCohortDataPreparations).

Centrally, the meta-analysis pipeline was run on the 52 cohorts. The pipeline which performs per cohort calculations of effect sizes and standard errors and the inverse variance meta-analysis is available at https://github.com/eQTLGen/MetaAnalysis. We included 4 genetic principal components, 20 gene expression principal components and other technical covariates (e.g. RNA integrity number) where available. Per every dataset, genes were included if the fraction of unique expression values was equal or greater than 0.8., Variants were included based on imputation quality, Hardy-Weinberg equilibrium (HWE) and minor allele frequency (MAF) (Mach R2 ≥ 0.4, HWE p ≥ 1×10-6 and MAF ≥ 0.01). For European ancestry datasets, allele frequency (AF) within 0.2 of the European reference AF). In an additional step, genes were filtered to include only those genes that were available in at least 50% of the cohorts and 50% of the RNA-seq samples. Variants were also filtered to those that were available in at least 50% of the cohorts.

### Author Information For eQTLGen Consortium

Habibul Ahsan1, Marta E. Alarcón-Riquelme2,3, Philip Awadalla4, Guillermo Barturen5,6,2, Alexis Battle7, Frank Beutner8, Cornelis Blauwendraat9,10, Collins Boahen11,12, Toni Boltz13, Dorret I. Boomsma14,15,16, Andrew Brown17, John Budde18,19, Katie L. Burnham20, John Chambers21,22,23, Evans Cheruiyot24, Surya B. Chhetri25, Annique Claringbould26,27, PRECISESADS Clinical Consortium2, DIRECT Consortium28, Carlos Cruchaga18,29,30,31,32,33, Kensuke Daida9,10,34, Emma E. Davenport20, Devin Dikec18,19, Théo Dupuis17, Diptavo Dutta35, Tõnu Esko36, Radi Farhad37, Aiman Farzeen38,39,40, Marie-Julie Favé41, Luigi Ferrucci42, Lude Franke26,43, Timothy M. Frayling44, Koichi Fukunaga45, J. Raphael Gibbs46, Greg Gibson47, Christian Gieger39,40, Priyanka Gorijala18,19, Marleen van Greevenbroek48, Binisha Hamal Mishra49,50,51, Takanori Hasegawa52, M. Arfan Ikram53, Michael Inouye54,55,56,57,58,59, Rick Jansen15,60, Farzana Jasmine1, Matt Johnson18,19, Mika Kähönen50,61, Muhammad G. Kibriya1, Holger Kirsten62,63, Julian C. Knight64, Peter Kovacs65,66, Knut Krohn67, Viktorija Kukushkina36, Vinod Kumar11,12,68,69, Sandra Lapinska70, Terho Lehtimäki49,50,51, Yun Li71,72,73, Markus Loeffler74,75, Marie Loh21,76,77,23, Leo-Pekka Lyytikäinen49,50,51, Reedik Mägi36, Javier Martin78, Angel Martinez-Perez79, Allan F. McRae24, Joyce van Meurs80,81, Lili Milani36, Pashupati P. Mishra49,50,51, Younes Mokrab37, Grant W. Montgomery24, Juha Mykkänen82,83, Haroon Naeem37, Sini Nagpal47, Ho Namkoong84, Matthias Nauck85, Yukinori Okada86,87,88,89,90, Roel Ophoff70,13,91, Katja Pahkala83,82,92, Bogdan Pasaniuc70,93,13,94, Dirk S. Paul54,55,95, Elodie Persyn54,55,96, Annette Peters40,97,98,99, Brandon Pierce1, René Pool100,15, Holger Prokisch101,38,102, Laura Raffield72, Venket Raghavan74, Olli T. Raitakari103,104,82,105, Emma Raitoharju106,107, María Rivas-Torrubia2, Ruth D. Rodríguez2, Suvi P. Rovio83,82, Jessie Sanford18,19, Markus Scholz74,75, Andrew Singleton108, Eline Slagboom109, José Manuel Soria79, Juan Carlos Souto79, Michael Stumvoll110,111,112, Yun Ju Sung18,19, Darwin Tay21, Alexander Teumer113,114,115, Joachim Thiery75,116, Alex Tokolyi20, Lin Tong1, Anke Tönjes110, Jan Veldink117, Joost Verlouw80, Ana Viñuela118, Peter M. Visscher24, Uwe Völker114,119, Urmo Võsa36, Qingbo S. Wang86,87,88, Robert Warmerdam26,43, Stefan Weiss120,114, Jia Wen72, Harm-Jan Westra26,43, Andrew R. Wood121, Manke Xie47, Dasha Zhernakova26

The author list is ordered alphabetically

1. Department of Public Health Sciences, University of Chicago, Illinois, USA

2. Pfizer–University of Granada–Junta de Andalucía Centre for Genomics and Oncological Research, Granada, Spain

3. Institute of Environmental Medicine, Karolinska Institute, Stockholm, Sweden

4. Ontario Institute for Cancer Research, University of Toronto

5. Department of Genetics, Faculty of Science, University of Granada, 18071 Granada, Spain

6. Bioinformatics Laboratory, Biotechnology Institute, Centro de Investigación Biomédica, PTS, Avda. del Conocimiento s/n, 18100 Granada, Spain

7. Department of Biomedical Engineering, Department of Computer Science, Department of Genetic Medicine, Johns Hopkins University, Baltimore, MD, USA

8. Department of Internal Medicine/Cardiology, Heart Center Leipzig at Leipzig University, Leipzig, Germany

9. Integrative Neurogenomics Unit, Laboratory of Neurogenetics, National Institute on Aging, National Institutes of Health, Bethesda, MD, USA

10. Center for Alzheimer’s and Related Dementias (CARD), National Institute on Aging and National Institute of Neurological Disorders and Stroke, National Institutes of Health, Bethesda, MD, USA

11. Department of Internal Medicine and Radboud Institute of Molecular Life Sciences (RIMLS), Radboud University Medical Center, Nijmegen, 6525 HP, the Netherlands

12. Department of Internal Medicine and Radboud Center for Infectious Diseases (RCI), Radboud University Medical Center, Nijmegen, 6525 HP, the Netherlands

13. Department of Human Genetics, David Geffen School of Medicine, University of California Los Angeles, Los Angeles, CA, USA

14. Amsterdam Reproduction & Development (AR&D) research institute, Amsterdam, the Netherlands

15. Amsterdam Public Health research institute, Amsterdam, the Netherlands

16. Department of Complex Trait Genetics, Center for Neurogenomics and Cognitive Research, Amsterdam, Vrije Universiteit Amsterdam

17. Population Health and Genomics, University of Dundee, Dundee, Scotland, UK.

18. Department of Psychiatry, Washington University School of Medicine, St. Louis, MO, United States

19. NeuroGenomics and Informatics Center, Washington University School of Medicine, St. Louis, MO 63110, USA

20. Wellcome Sanger Institute, Wellcome Genome Campus, Hinxton, UK

21. Lee Kong Chian School of Medicine, Nanyang Technological University, Singapore

22. Precision Health Research (PRECISE), Singapore

23. Department of Epidemiology and Biostatistics, School of Public Health, Imperial College London, United Kingdom

24. Institute for Molecular Bioscience, The University of Queensland, Brisbane, Australia

25. Department of Biomedical Engineering, Johns Hopkins University, Baltimore, MD, USA

26. Department of Genetics, University Medical Center Groningen, University of Groningen, Groningen, The Netherlands

27. Department of Internal Medicine, Erasmus MC, Erasmus University Medical Center Rotterdam, Rotterdam, The Netherlands

28. https://directdiabetes.org/

29. NeuroGenomics and Informatics Center, Washington University School of Medicine, St. Louis, MO 63110, USA

30. Department of Neurology, Washington University School of Medicine, St. Louis, MO 63110, USA

31. Knight Alzheimer Disease Research Center, Washington University School of Medicine, St. Louis, MO, United States

32. Hope Center for Neurological Disorders, Washington University School of Medicine, St. Louis, MO, United States

33. Dominantly Inherited Alzheimer Disease Network (DIAN)

34. Department of Neurology, Faculty of Medicine, Juntendo University, Tokyo, Japan

35. Division of Cancer Epidemiology & Genetics, National Cancer Insititute, Bethesda, MD, USA

36. Estonian Genome Centre, Institute of Genomics, University of Tartu, Tartu, Estonia

37. Human Genetics Department, Sidra Medicine, Doha, Qatar

38. Institute of Neurogenomics, Computational Health Center, Helmholtz Munich, Neuherberg, Germany

39. Research Unit of Molecular Epidemiology, Helmholtz Zentrum München - German Research Center for Environmental Health, Neuherberg, Germany

40. Institute of Epidemiology, Helmholtz Zentrum München - German Research Center for Environmental Health, Neuherberg, Germany

41. Ontario Institute for Cancer Research

42. Translational Gerontology Branch, National Institute on Aging, National Institutes of Health, Baltimore, MD, USA

43. Oncode Institute, Groningen, The Netherlands

44. Department of Genetic Medicine and Development, CMU, University of Geneva

45. Division of Pulmonary Medicine, Department of Medicine, Keio University School of Medicine, Tokyo, Japan.

46. Computational Biology Group, Laboratory of Neurogenetics, National Institute on Aging, Bethesda, MD, USA

47. Center for Integrative Genomics, Georgia Institute of Technology, Atlanta, GA, USA.

48. CARIM, Maastricht University

49. Department of Clinical Chemistry, Faculty of Medicine and Health Technology, Tampere University, Tampere, Finland

50. Finnish Cardiovascular Research Center Tampere, Faculty of Medicine and Health Technology, Tampere University, Tampere, Finland

51. Department of Clinical Chemistry, Fimlab Laboratories, Tampere, Finland

52. M&D Data Science Center, Tokyo Medical and Dental University, Tokyo, Japan.

53. Department of Epidemiology, Erasmus MC University Medical Center, Rotterdam, The Netherlands

54. British Heart Foundation Cardiovascular Epidemiology Unit, Department of Public Health and Primary Care, University of Cambridge, Cambridge, UK

55. Victor Phillip Dahdaleh Heart and Lung Research Institute, University of Cambridge, Cambridge, UK

56. Cambridge Baker Systems Genomics Initiative, Department of Public Health and Primary Care, University of Cambridge, Cambridge, UK

57. Cambridge Baker Systems Genomics Initiative, Baker Heart and Diabetes Institute, Melbourne, VIC, Australia

58. Health Data Research UK Cambridge, Wellcome Genome Campus and University of Cambridge, Cambridge, UK

59. British Heart Foundation Centre of Research Excellence, University of Cambridge, Cambridge, UK.

60. Amsterdam UMC location Vrije Universiteit Amsterdam, Department of Psychiatry & Amsterdam Neuroscience -Complex Trait Genetics (VUmc) and Mood, Anxiety, Psychosis, Stress & Sleep

61. Department of Clinical Physiology, Tampere University Hospital, Tampere Finland

62. LIFE – Leipzig Research Center for Civilization Diseases, Leipzig University, Germany

63. Institute for Medical Informatics, Statistics and Epidemiology, Leipzig University, Germany

64. Centre for Human Genetics, University of Oxford, Oxford, UK

65. Integrated Research and Treatment Center (IFB) Adiposity Diseases, University of Leipzig, D-04103 Leipzig, Germany

66. Department of Medicine, University of Leipzig, D-04103 Leipzig, Germany

67. Medical Faculty, University of Leipzig, Leipzig, Germany.

68. Department of Genetics, University of Groningen, University Medical Center Groningen, Groningen, 9700 RB, the Netherlands

69. Nitte (Deemed to Be University), Medical Sciences Complex, Nitte University Centre for Science Education and Research (NUCSER), Deralakatte, Mangalore, 575018, India

70. Bioinformatics Interdepartmental Program, University of California Los Angeles, Los Angeles, CA, USA

71. Department of Biostatistics, University of North Carolina at Chapel Hill, Chapel Hill, NC, USA

72. Department of Genetics, University of North Carolina, Chapel Hill, NC, USA

73. Department of Computer Science, University of North Carolina at Chapel Hill, Chapel Hill, NC, USA

74. Institute for Medical Informatics, Statistics and Epidemiology, Leipzig University, Leipzig, Germany

75. Leipzig Research Centre for Civilization Diseases, Leipzig University, Leipzig, Germany

76. National Skin Centre, Research Division, Singapore

77. Genome Institute of Singapore, Agency for Science, Technology and Research, Singapore

78. Instituto de Parasitología y Biomedicina López-Neyra, Consejo Superior de Investigaciones Científicas (IPBLN-CSIC), Granada, Spain

79. Unit of Genomic of Complex Diseases, Institut de Recerca Sant Pau (IR Sant Pau), Barcelona, Spain.

80. Department of Internal Medicine, Erasmus MC University Medical Center, Rotterdam, The Netherlands

81. Department of Orthopaedics and Sportsmedicine, Erasmus MC University Medical Center, Rotterdam, The Netherlands.

82. Centre for Population Health Research, University of Turku and Turku University Hospital, Turku, Finland

83. Research Centre of Applied and Preventive Cardiovascular Medicine, University of Turku, Turku, Finland

84. Department of Infectious Diseases, Keio University School of Medicine, Tokyo, Japan.

85. Institute of Clinical Chemistry and Laboratory Medicine, University Medicine Greifswald, Greifswald, Germany

86. Department of Genome Informatics, Graduate School of Medicine, the University of Tokyo, Tokyo, Japan

87. Department of Statistical Genetics, Osaka University Graduate School of Medicine, Suita, Japan

88. Laboratory for Systems Genetics, RIKEN Center for Integrative Medical Sciences, Yokohama, Japan

89. Laboratory of Statistical Immunology, Immunology Frontier Research Center (WPI-IFReC), Osaka University, Suita, Japan

90. Premium Research Institute for Human Metaverse Medicine (WPI-PRIMe), Osaka University, Suita, Japan.

91. Center for Neurobehavioral Genetics, Semel Institute for Neuroscience and Human Behavior, David Geffen School of Medicine, University of California Los Angeles, Los Angeles, USA

92. Paavo Nurmi Centre, Unit of Health and Physical Activity, University of Turku, Turku, Finland

93. Department of Computational Medicine, David Geffen School of Medicine, University of California Los Angeles, Los Angeles, CA, USA

94. Department of Pathology and Laboratory Medicine, David Geffen School of Medicine, University of California Los Angeles, Los Angeles, CA, USA

95. Centre for Genomics Research, Discovery Sciences, BioPharmaceuticals R&D, AstraZeneca, Cambridge, UK.

96. Cambridge Baker Systems Genomics Initiative, Department of Public Health and Primary Care, University of Cambridge, UK.

97. Chair of Epidemiology, IBE, Faculty of Medicine, LMU Munich, Munich, Germany

98. German Centre for Cardiovascular Research (DZHK), Partner Site Munich Heart Alliance, Munich, Germany

99. German Center for Diabetes Research (DZD), Neuherberg, Germany.

100. Department of Biological Psychology, Vrije Universiteit Amsterdam, Amsterdam, the Netherlands

101. School of Medicine, Institute of Human Genetics, Technical University of Munich, Munich, Germany

102. German Center for Child and Adolescent Health (DZKJ), partner site Munich, Munich, Germany

103. Research centre of Applied and Preventive Cardiovascular Medicine, University of Turku, Turku, Finland

104. Department of Clinical Physiology and Nuclear Medicine, Turku University Hospital, Turku, Finland

105. InFLAMES Research Flagship, University of Turku, Turku, Finland

106. Molecular Epidemiology, Faculty of Medicine and Health Technology, Tampere University, Tampere, Finland

107. Tampere University Hospital, Tampere, Finland

108. Laboratory of Neurogenetics, National Institute on Aging, National Institutes of Health, Bethesda, MD, USA

109. Section of Molecular Epidemiology, Department of Biomedical Data Sciences, Leiden University Medical Center, Leiden, the Netherlands

110. Medical Department III-Endocrinology, Nephrology, Rheumatology, University of Leipzig Medical Center, Leipzig, Germany

111. Helmholtz Institute for Metabolic, Obesity and Vascular Research (HI-MAG), Helmholtz Zentrum München, University of Leipzig and University Hospital Leipzig, Leipzig, Germany

112. Deutsches Zentrum für Diabetesforschung, Neuherberg, Germany

113. Department of Psychiatry and Psychotherapy, University Medicine Greifswald, Greifswald, Germany

114. DZHK (German Center for Cardiovascular Research), Partner Site Greifswald, Greifswald, Germany

115. Department of Population Medicine and Lifestyle Diseases Prevention, Medical University of Bialystok, Bialystok, Poland

116. Institute of Laboratory Medicine, Clinical Chemistry and Molecular Diagnostics, University of Leipzig Medical Center, Leipzig, Germany

117. Department of Neurology, UMC Utrecht Brain Center Rudolf Magnus, Utrecht, The Netherlands

118. Biosciences Institute, Faculty of Medical Sciences, University of Newcastle, Newcastle upon Tyne, UK

119. Interfaculty Institute for Genetics and Functional Genomics, University Medicine Greifswald, Felix-Hausdorff-Strasse 8, D-17475 Greifswald, Germany

120. Interfaculty Institute of Genetics and Functional Genomics, University Medicine Greifswald, Greifswald, 17475, Germany

121. College of Medicine and Health, University of Exeter, Exeter, UK
